# Wastewater surveillance suggests unreported Mpox cases in a low-prevalence area

**DOI:** 10.1101/2023.05.28.23290658

**Authors:** Jeremiah Oghuan, Carlos Chavarria, Scout R. Vanderwal, Anna Gitter, Akpevwe Amanda Ojaruega, Carlos Monserrat, Cici X. Bauer, Eric L. Brown, Sara Javornik Cregeen, Jennifer Deegan, Blake M. Hanson, Michael Tisza, Hector I Ocaranza, John Balliew, Anthony W. Maresso, Janelle Rios, Eric Boerwinkle, Kristina D. Mena, Fuqing Wu

## Abstract

Wastewater surveillance has emerged as a valuable tool for monitoring infectious disease agents including SARS-CoV-2 and Mpox virus. However, detecting the Mpox virus in wastewater is particularly challenging due to its relatively low prevalence in the community. In this study, we detected Mpox virus in wastewater from a US-Mexico border city with a low prevalence of Mpox disease during February and March 2023 using real-time PCR assays targeting the C22L, F3L, and F8L genes. An increasing trend of viral concentration was observed 1∼2 weeks earlier than when the Mpox case was reported. Further sequencing and epidemiological analysis provided supporting evidence for unreported Mpox infections in the city. This study showcases a combined approach with multiple molecular assays for efficient detection of the Mpox virus in wastewater in a low-prevalence area. The findings emphasize the value of wastewater surveillance as a timely identification tool for infectious diseases in low-prevalence areas, and the need for heightened vigilance to control the spread of infectious diseases in such settings.

## Introduction

The Mpox (formerly referred to as monkeypox) outbreak in 2022 was a significant public health event. The Mpox virus is a linear, double-stranded DNA virus in the genus *Orthopoxvirus*. The Mpox virus (MPXV) was first isolated from a sick monkey in 1959 ^1^ and the first human infection was confirmed in 1970 in a 9-month-old newborn in the Democratic Republic of the Congo ^2^. As a zoonotic disease, Mpox can be transmitted through intimate contact with respiratory secretions, skin lesions of an infected person, and exposure to blood, body fluids, cutaneous or mucosal lesions from infected animals ^3^. The World Health Organization declared the rapid spread of Mpox as a public health emergency of international concern on July 23, 2022. As of May 2023, there have been more than 3,000 cases reported in Texas, and globally 86,000 confirmed cases in 110 countries, of which 103 had no historically reported Mpox infections ^4^.

Wastewater surveillance is a promising approach for monitoring community outbreaks of infectious diseases. Wastewater collates viral signals excreted by infected individuals across the whole spectrum of disease symptoms ranging from asymptomatic to pre-symptomatic to symptomatic ^5^. This inclusiveness of all virus-carrying individuals in the sewershed enables us to track the epidemic progression and better estimate the magnitude of infections^6,7^. In addition, wastewater surveillance can capture the shedding of viral particles prior to the onset of symptoms providing an early warning of emerging outbreaks in the population ^8–10^. For example, Wolfe *et al*. reported MPXV DNA detection in multiple wastewater sampling sites in California and some sites detected Mpox before identification of cases^11^.

The Centers for Disease Control and Prevention (CDC) recommended two real-time PCR assays for the detection of MPXV (C22L) ^12^ and non-variola orthopoxvirus (F8L) ^13^ in June 2022. We previously confirmed that the sequence of primers and probes in the two assays matches >99.4% and >99.1% of global Mpox genomes, respectively ^14^. Two nucleotide mismatches were also found in the primers of C22L assay, and mismatch-corrected primers (C22L_m) showed an improved detection sensitivity with the 100% limit of detection of 2.7 copies of DNA per reaction. We also identified that an additional assay targeting MPXV F3L gene ^15^ has a high homology to >99.7% of Mpox genomes. In this study, we further characterized their analytical sensitivity and limit of detection and used the three assays (F3L, F8L, and C22L_m) to detect and track MPXV in wastewater samples collected from four wastewater treatment plants serving the border city of El Paso, Texas during February and March in 2023.

## Materials and Methods

### Wastewater sample collection and processing

Weekly, 24-h composite wastewater samples were collected from the City of El Paso, Texas from February 20 to March 27, 2023. There are four wastewater treatment facilities (abbreviated names: FH, HS, JT, and RB) serving a total of 751,982 individuals in the city. Samples were collected by El Paso Water Utility and shipped overnight on ice to the laboratory at Houston for analysis. Samples were processed on the day of receipt. Raw wastewater samples were separated into pellet and supernatant portions. Specifically, 2 mL of raw wastewater was aliquoted and centrifuged to collect the pellet for DNA extraction. For supernatants, 40 mL raw wastewater samples were vacuum filtered through a 0.22 μm polyether sulfone membrane to remove cell debris and solid materials. Supernatants were used to concentrate viral particles as described below.

### Viral concentration and DNA extraction

Viral enrichment was performed based on previous methods^8,16,17^. Briefly, 15 ml of filtrates were concentrated with 30 kDa Amicon Ultra Centrifugal Filter (Sigma, Cat#: UFC9010) by centrifugation (3,900 rpm for 20 mins) to 150 -200 μl subsequently used for DNA extraction using QIAamp DNA Mini Kit (Qiagen, Cat#: 51306). The concentrates were resuspended with 200 μl buffer ATL for lysis. For pellet, ATL buffer was directly added into the pellet for DNA extraction. Proteinase K was added into the sample to facilitate the lysis followed by 2 h incubation at 56°C. DNA extraction was performed based on the standard procedures provided by the kit and 100 μl nuclease-free H_2_O was used to elute DNA extracted from pellets and supernatants.

### Real-Time quantitative PCR (RT-qPCR)

The eluted DNA was used to test MPXV by probe-based RT-qPCR. Briefly, PCR Multiplex Supermix (Bio-Rad, Cat #: 12010220) was mixed with the primers, probe, and nuclease-free H_2_O (VWR, Cat #: 10220-402) and then added to respective wells of a 96-well PCR plate (Bio-Rad, Cat #: HSL9605). The final concentration for the primers and probe is 750 nM and 375 nM, respectively. The reaction was run with the following program: 95°C for 3 minutes for denature, followed by 48 cycles of denature (95 °C 3 s) and anneal/extend (60 °C 30 s). All primers, probes, and synthetic gene fragments for MPXV, cowpox, and horsepox viruses were synthesized from Integrated DNA Technologies and their sequence was provided in **Table S3**. Detailed procedures for the RT-qPCR can be found in our previous work ^14^. The standard curves for F3L and F8L using different DNA templates were generated by serial ten-fold dilutions of the synthesized gene fragment from 3.64∼3.64e6 copies/µl. Each concentration has 6 technical replicates, and the mean value with standard deviation is used for plotting. Two-fold serial dilutions were further used to determine the limit of determination (LOD) for both assays with different DNA templates. In total, 16∼22 replicates were performed at critical concentrations. The cycle threshold (*C*_t_) value was exported by the built-in software with manual confirmation of a sigmoidal amplification curve.

For wastewater samples, we used the F3L, F8L, and C22L_m assays to test MPXV in the supernatant and pellet of each sample. Three technical PCR replicates were performed for each assay. Negative controls (i.e., adding the same volume of nuclease-free H2O with no DNA template) were included for each running with a total of 16∼24 replicates. To minimize the potential cross-contamination, we added the DNA for positive controls after all the other samples were prepared and before the sealing of the 96-well plate, followed by centrifugation at 1,000 rpm for 1 min before the real-time PCR running. For all the real-time PCR results, *C*t value above 42 was considered negative.

Sanger sequencing was performed to verify the amplified sequence. Specifically, we used the forward and reverse primers in the C22L_m assay (Table S3) to amplify wastewater samples’ DNA using Ultra II Q5 Master Mix (New England BioLabs, Cat#: M0544L). PCR products were then sent to Eton Bioscience Inc. for Sanger sequencing using the C22L_m-forward primer. The size of C22L_m amplicon is 90 bp.

### Sequence alignment and mismatch analysis

Complete genome sequences were downloaded from National Center for Biotechnology Information (NCBI) as fasta files, including 3,282 genomes for human Mpox viruses, 98 Cowpox viruses, 4 Horsepox viruses, 58 Smallpox viruses, 112 Vaccinia viruses, and 6 Buffalopox viruses released in the NCBI database as of February 13, 2023. The sequence was imported into R (version 4.1.3) and combined as the genome database for analysis. We aligned each of the oligo**s** (including forward primer, reverse primer, and probe) sequences and their reverse complements to the database and computed the percentage of genomes where the oligo sequences were a 100% match. For non-matching oligos, sequences were extracted and aligned using SnapGene (https://www.snapgene.com/) to identify the specific nucleotides that differ between the oligos and the genomic data. The major types of sequence variation(s) are listed in Figure 1 and Table S2. These analyses were performed with customized R scripts.

**Figure 1.**
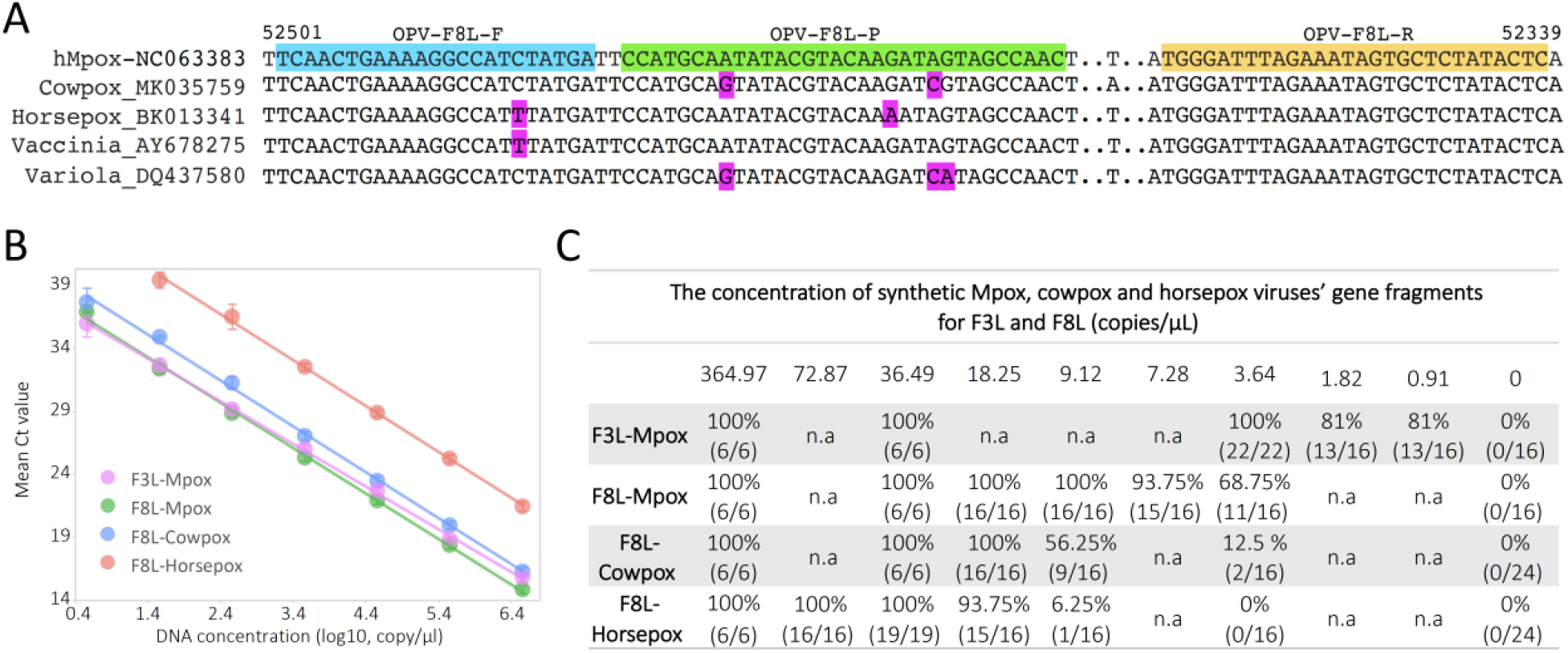
Specificity, sensitivity, and limit of detection for the F3L and F8L assays. (**A**) Alignment of F8L forward primer (highlighted cyan), probe (green), and reverse primer (yellow) against MPXV, cowpox viruses, horsepox, vaccinia virus, and smallpox (variola) virus genomes. Mismatched nucleotides were highlighted in red. (**B**) Standard curves for the F3L and F8L assays for Mpox, cowpox, and horsepox virus DNA detection. The data shown represent the mean of 6 replicates with standard deviations. Trendline equations and coefficients of determination are provided in Table S1. (**C**) Limits of detection of the F3L and F8L assays for Mpox, cowpox, and horsepox DNA fragments. n.a represents ‘not tested’. The number of replicates is shown in parentheses.

## Results

We first evaluated the analytical sensitivity and limit of detection of F3L and F8L assays using real-time quantitative PCR (RT-qPCR) with synthetic gene fragments covering corresponding genomic regions of MPXV and other *orthopoxvirus* viruses (**Fig. 1**). By testing the F3L assay using serial tenfold dilutions of MPXV DNA, we found that F3L detected MPXV DNA across 7-log concentrations from 3.64∼3.64*10^6^ copies/µl with an amplification efficiency of 97.62% and R^2^ > 0.999 (**Fig. 1B and Table S1**). The limit of detection with 22 replicates shows that the F3L assay is 100% sensitive to MPXV detection at 3.64 copies/µl (7.28 copies/reaction, **Fig. 1C**). Sixteen replicates of controls with no DNA template are negative.

The F8L assay was designed for broad detection of viruses in the genus *orthopoxvirus*; however, the homology between the sequence of primers/probe and species in the genus varies (**Fig. 1A**). We therefore aligned the sequence of primers and probe of the F8L assay against 6 *orthopoxvirus* species including 3,282 genomes for human Mpox viruses, 98 cowpox viruses, 4 horsepox viruses, 58 smallpox viruses, 112 vaccinia viruses, and 6 buffalopox viruses released in the NCBI database as of February 13, 2023. Overall, the F8L primers and probe have the highest full match to MPXV genomes (99.8%) and have 1∼3 nucleotide mismatches to most of the genomes (52%∼100%) in other species (**Table S2**), suggesting that the F8L assay may have varied detection sensitivity to different *orthopoxvirus* species.

To determine how these sequence variations impact detection of orthopoxvirus, we synthesized MPXV, cowpox, and horsepox virus gene fragments covering the F8L region. Standard curves from serial 7-log dilutions show similar amplification efficiencies using MPXV (89.49%), cowpox (88.40%), and horsepox DNA (88.39%) with R^2^ > 0.998 (**Fig. 1B and Table S1**). However, the analytical sensitivity ranges between 1.8 and 7.1 Ct among the three viral genome fragments. The 100% of detection of the three viral DNA sequences varies from 9.1 to 36.44 copies/µl (**Fig. 1C**). In total, 24 no-template controls for the F8L assay are negative. These results showed that the F8L assay is most sensitive in detecting this MPXV genome fragment compared to cowpox and horsepox viruses.

Next, we tested the detection of MPXV in wastewater using F3L, F8L, and C22L_m assays. Weekly wastewater samples were collected between February 20 and March 27, 2023 from four wastewater treatment facilities including FH, HS, JT, and RB, which together serve about 751,982 individuals in El Paso ^18^. Raw wastewater samples were separated into supernatant (filtrate) and pellet portions, which were individually tested by the three assays. Positive MPXV signals were observed in the supernatant or pellet or both (**Fig. S1**). Sanger sequencing of PCR products using FH and HS pellet samples on March 13 further confirmed 86% to 94% identity matches to the MPXV reference genomes (**Fig. S2**). The F3L assay detected MPXV in 17/24 (70.8%) samples, C22L_m detected the virus in 14/24 (58.3%) samples, and F8L in 3/24 (12.5%) samples (**Fig. 2A**). Eleven of 24 (52.4%) samples were detected by more than one assay, and three samples were not detected by any of the three assays. The HS sewershed had the highest F3L assay positivity frequency. Aggregating the viral concentrations by sampling date, viral concentrations in the pellet were relatively higher than in the supernatant (**Fig. 2B and Fig. S1**). Increased viral concentrations in wastewater were observed from February 27, with a peak in the week of March 6.

**Figure 2.**
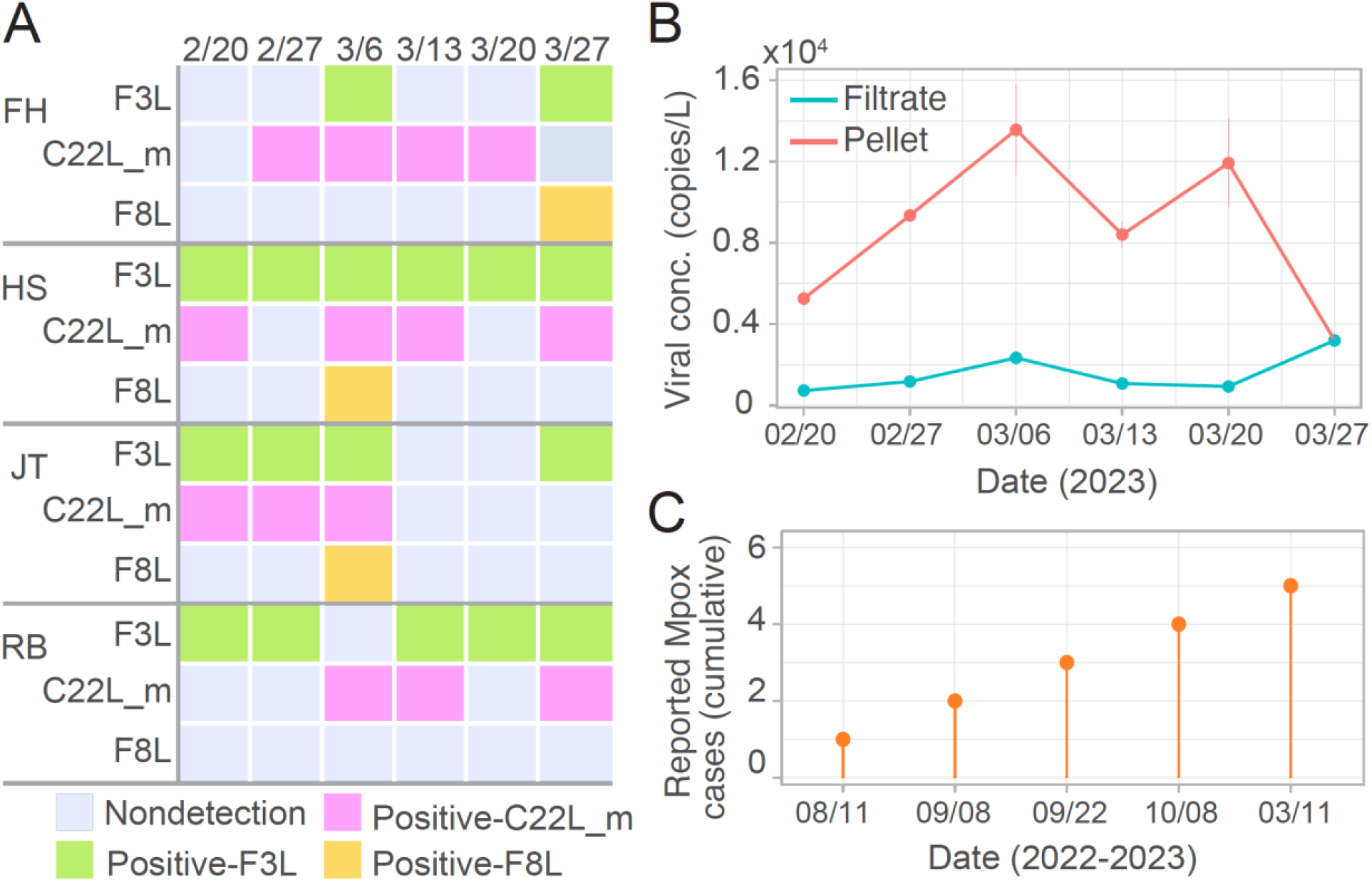
MPXV detection in wastewater using the F3L, F8L, and C22L_m assays. (**A**) Detection results using the F3L, F8L, and mismatch-corrected CDC (C22L_m) assays for weekly wastewater samples collected between February 20 and March 27, 2023. (**B**) Mean viral concentrations in wastewater supernatants and pellets. Data are expressed as the mean of four sewersheds and the three assays. Error bars represent the standard error of the mean. (**C**) Reported Mpox cases in the County. The most recent case was reported on March 11, 2023.

Clinically, five Mpox cases were reported since August 2022, and the most recent case in the city was found on March 11 (**Fig. 2C**). The time difference between wastewater peak time and new case reporting dates highlights the feasibility and precedence of wastewater surveillance for detecting new infections, also revealed by a recent study^11^. Positive signals were still observed in the four sewersheds after the case was identified and isolated (**Fig. S1**) suggesting that there might be additional unreported Mpox cases. Nucleotide variations of the C22L_m amplicons’ sequence for samples collected from sewersheds FH and HS on March 13 (**Fig. S2**) supports the hypothesis. This is further supported by following findings: a) positive signals were found on February 20, approximately three weeks before the latest case was reported which exceeds the mean viral incubation period of 5.6 to 9.1 days^19,20^ and b) the penultimate case was reported on October 8, 2022. Persistent viral shedding in patients has been reported for up to 39 days in the literature^21,22^, but has never been reported for over 4 months, suggesting that the signals in wastewater on February 20 may not be from shedding from October 8 case. These analyses indicated the presence of unreported Mpox cases in the city.

## Discussion

Wastewater surveillance is being widely implemented as a complementary public health tool to detect infectious pathogens including SARS-CoV-2 and Mpox virus^11,23–25^. However, it is more challenging to detect the Mpox virus in wastewater because of its relatively lower prevalence in the community leading to lower viral concentrations. The presence of inhibitors in wastewater samples further challenges the efficiency of molecular detection approaches. Using multiple MPXV detection assays can partially address this challenge. Taking the C22L_m assay in HS sewershed as an example, we did not detect MPXV DNA in either the supernatant or pellet for samples collected on February 27 and March 20; however, other samples collected during this time frame were positive. There are many potential reasons for the negative tests including low viral DNA concentrations and PCR inhibitors present in wastewater. As such, results using the F3L and F8L assays were crucial for cross-validation. Thus, using multiple assays targeting different genomic sites of the virus can enhance detection accuracy, especially in areas with low prevalence.

While Mpox cases continue to be reported globally, the percentage of asymptomatic and or unreported MPXV infections remains unknown. Two recent studies found 75%^26^ and 5%^27^ of asymptomatic infections among 4 and 284 Mpox cases by screening 224 and 583 individuals, respectively. Although the range is large, both studies showed that the current number of Mpox cases is underestimated, which is further supported by our wastewater results reported here. The next important question that arises is how to effectively identify the specific locations of these non-reported infections within the city. To address this, one potential approach is to strategically sample wastewater from upstream communities, such as neighborhoods, housing estates, and even individual buildings ^28–30^. Analyzing these upstream wastewater samples can aid in pinpointing the locations with non-reported infections within the city, and facilitate targeted public health interventions.

In summary, we detected MPXV using three distinct molecular assays in wastewater samples collected from El Paso, Texas. Notably, we observed a progressive rise in viral concentrations in the wastewater approximately 1-2 weeks prior to the reporting of a new clinical case. The presence of viral signals in multiple sewersheds both before and after the identification of the sole clinical case strongly suggests the existence of unreported Mpox virus infections within the city during the sampling period. This study emphasizes the importance of employing a combined approach that incorporates multiple well-established molecular assays to improve the detection of Mpox virus DNA in wastewater, especially in regions with a low prevalence of the disease.

## Data Availability

All data produced in the present study are available upon reasonable request to the authors.

## Code Availability

All data and code produced in the present study are available upon reasonable request to the authors.

## Acknowledgment

We thank Teresa T. Alcala and Xavier Soto at El Paso Water, and Camille J. Breaux and Malini Udtha at UTHealth Houston for sample coordination, collection, and shipping. We are grateful to Carolyn S. Wade, David W. Jackson, and Linda Bowen at UTHealth Houston for ordering materials and administration. This work is supported by Faculty Startup funding from the Center of Infectious Diseases at UTHealth, the UT system Rising STARs award, and the Texas Epidemic Public Health Institute (TEPHI).

## Author Contribution

Conceptualization: KDM, FW; Investigation: JO, FW; Methodology: FW; Visualization: JO, FW; Supervision: KDM, FW; Funding acquisition, JR, EB, KDM, FW; Discussion: JO, CC, SRV, AG, AAO, CM, CXB, ELB, SJC, JD, BMH, MT, HIO, JB, AWM, JR, EB, KDM, FW; Writing – original draft: JO, FW; Writing – review & editing: JO, CC, SRV, AG, AAO, CM, CXB, ELB, SJC, JD, BMH, MT, HIO, JB, AWM, JR, EB, KDM, FW.

## Ethical Approval Statement

This article does not contain any studies involving animals or humans performed by any authors.

## Supplementary Data

**Figure S1.**
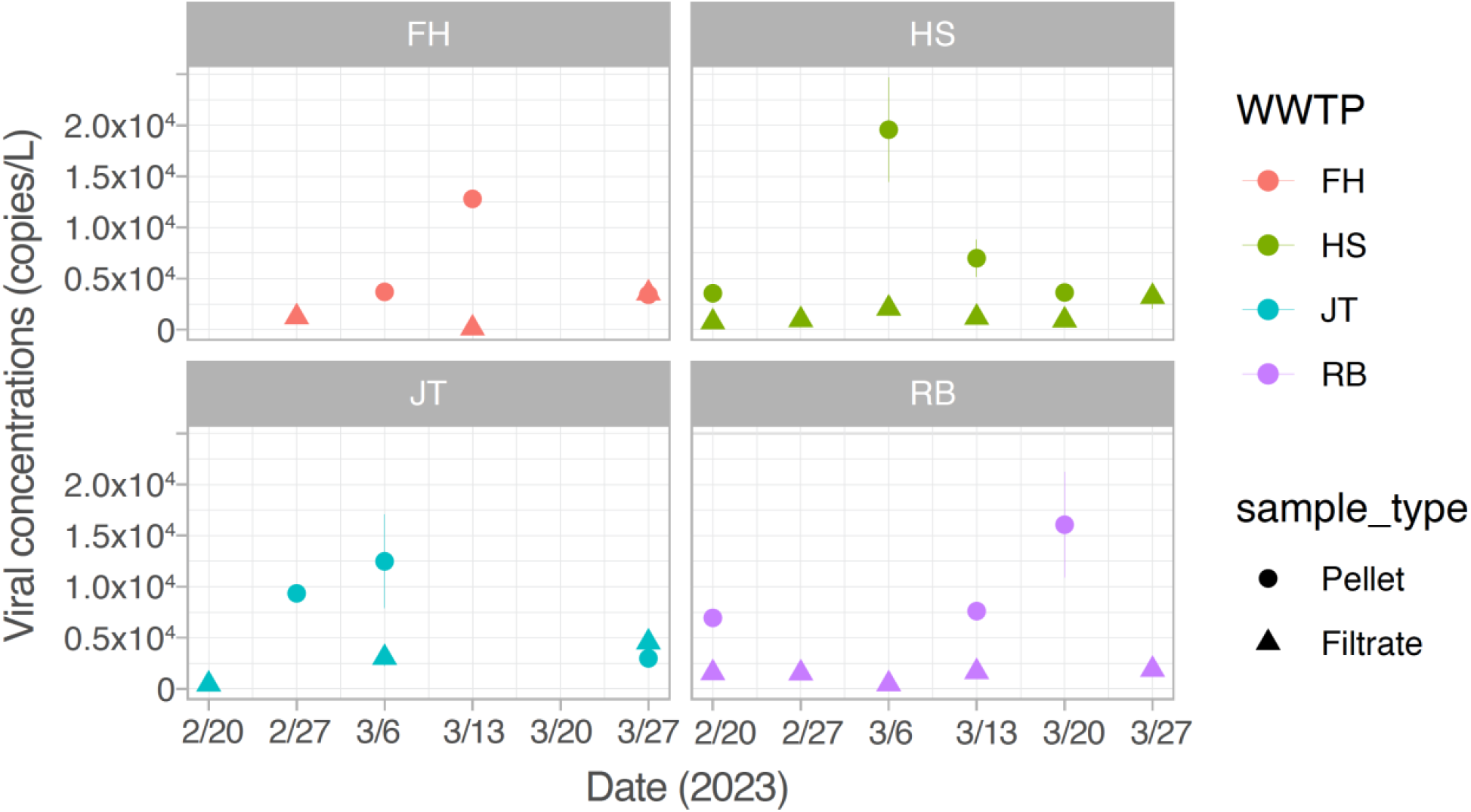
Mean viral concentrations by sampling date in the four sewersheds. Data were aggregated by the three tested assays and only positive data points are plotted. Error bars represent the standard error of the mean of results from the 3 assays each with 3 technical replicates.

**Fig. S2.**
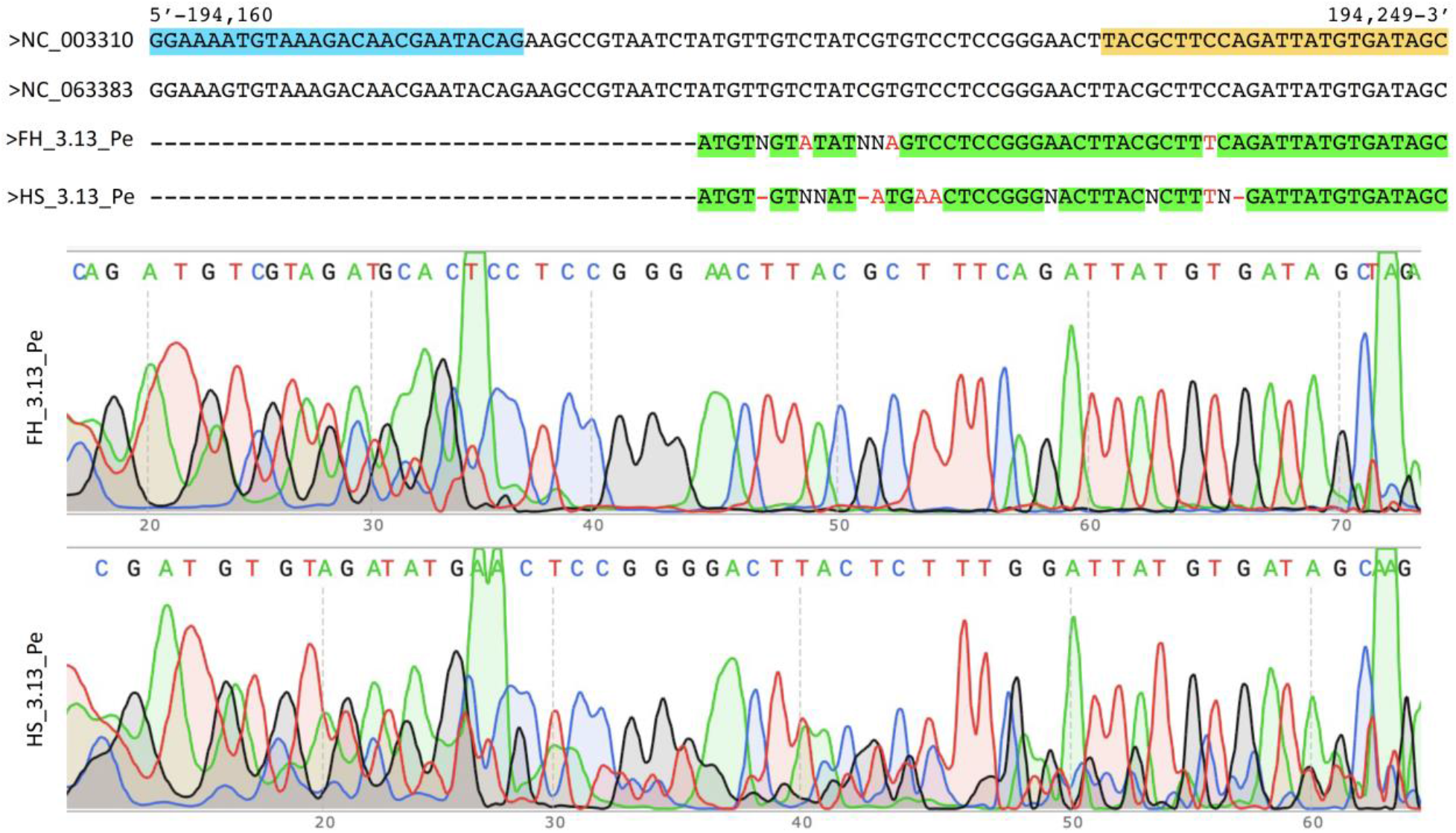
Sanger sequencing result and alignment to MPXV genome. Top: the sequence of PCR products by C22L_m assay was aligned to MPXV reference genomes (GenBank accession number: NC_003310 and NC_063383). The amplicon size of the C22L_m assay is 90 bp. The amplification was using wastewater pellet samples from FH and HS on March 13, 2023. Highlighted green: aligned sequence between PCR products and reference genomes; colored red: unaligned nucleotides; highlighted cyan: forward primer region; highlighted yellow: reverse primer region. Ambiguous base pairs were labeled as “N”. Bottom: sequencing chromatograms for the PCR products (FH and HS) using forward primer.

**Table S1.** Standard curves for F3L and F8L assays using Mpox, cowpox, and horsepox virus genome fragments.

| RT-qPCR assays | y-intercept | Slope of the standard curve | Amplification efficiency | Coefficient of determination (R<sup>2</sup>) |
| --- | --- | --- | --- | --- |
| F3L-Mpox | 37.91 | -3.3804 | 97.62% | 0.9996 |
| F8L-Mpox | 38.333 | -3.6025 | 89.49% | 0.9984 |
| F8L-Cowpox | 40.172 | -3.6352 | 88.40% | 0.9984 |
| F8L-Horsepox | 45.428 | -3.6357 | 88.39% | 0.9987 |

Note: x-axis: DNA concentrations (log10, copy/μl); y-axis: Ct values.

**Table S2.** Summary of the alignment of F8L oligos to the 6 species in *orthopoxvirus*.

| OPV-F8L oligos | hMpox (3282)<sup>1</sup> | Cowpox virus (98) | Horsepox (4) | Smallpox (58) | Vaccinia virus (112) | Buffalopox (6) |
| --- | --- | --- | --- | --- | --- | --- |
| OPV-F8L-F | 3277 Matches | 96 Matches | 4 Mismatches (Vaccinia_AY678275) | 56 Matches | 81 Matches; 30 Mismatches (Vaccinia_AY678275) | 5 Mismatches (Vaccinia_AY678275) |
| OPV-F8L-R | 3276 Matches | 79 Matches | 4 Matches | 57 Matches | 110 Matches | 6 Matches |
| OPV-F8L-P | 3276 Matches | 42 Matches; 51 Mismatches (Cowpox_MK035759) | 4 Mismatches (Horsepox_BK013341) | 57 Mismatches (Variola_DQ437580) | 91 Matches; 19 Mismatches (Horsepox_BK013341) | 6 Matches |

<sup>1</sup> The number of complete genomes in NCBI as of February 13, 2023. hMpox: human monkeypox virus.

**Table S3.**
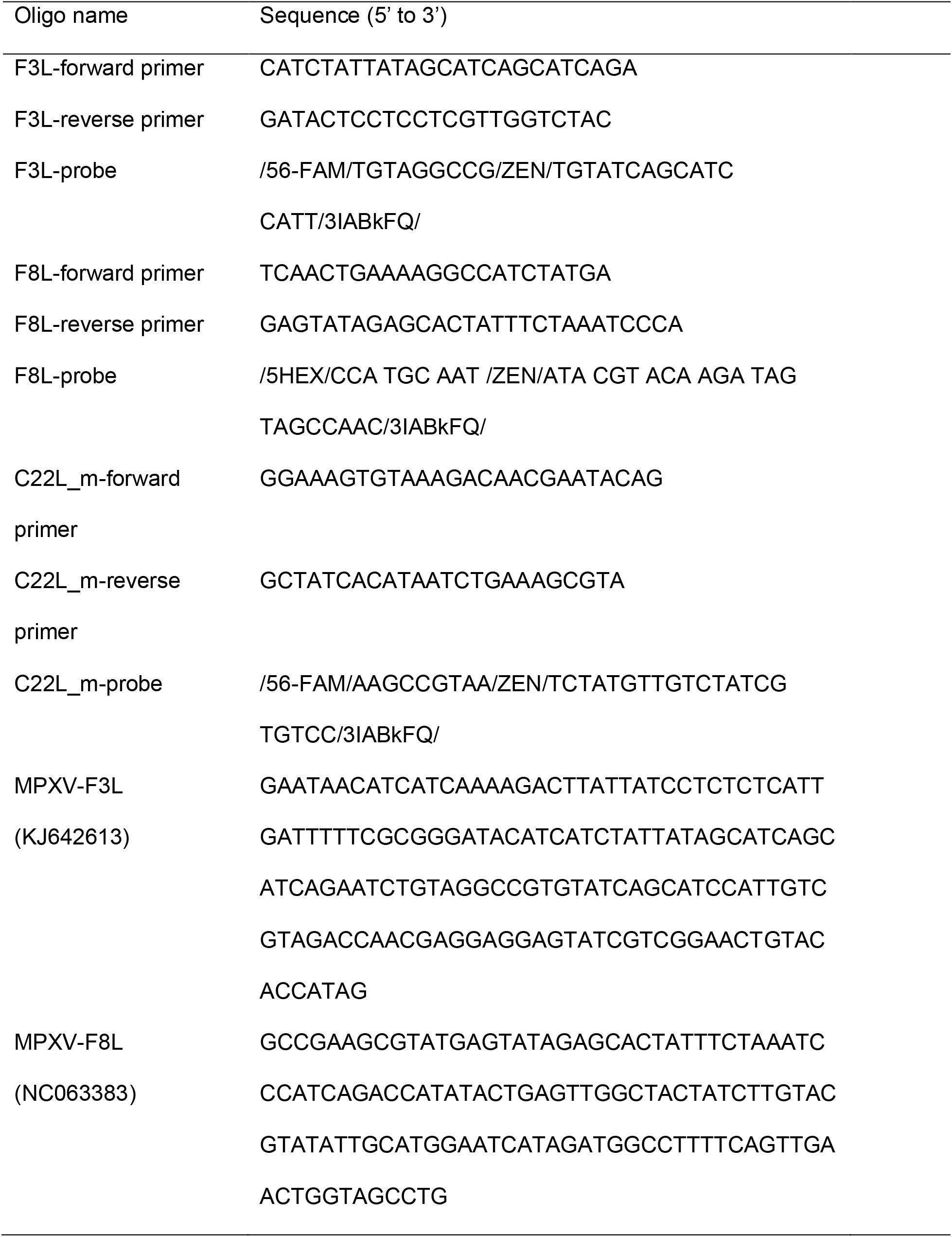
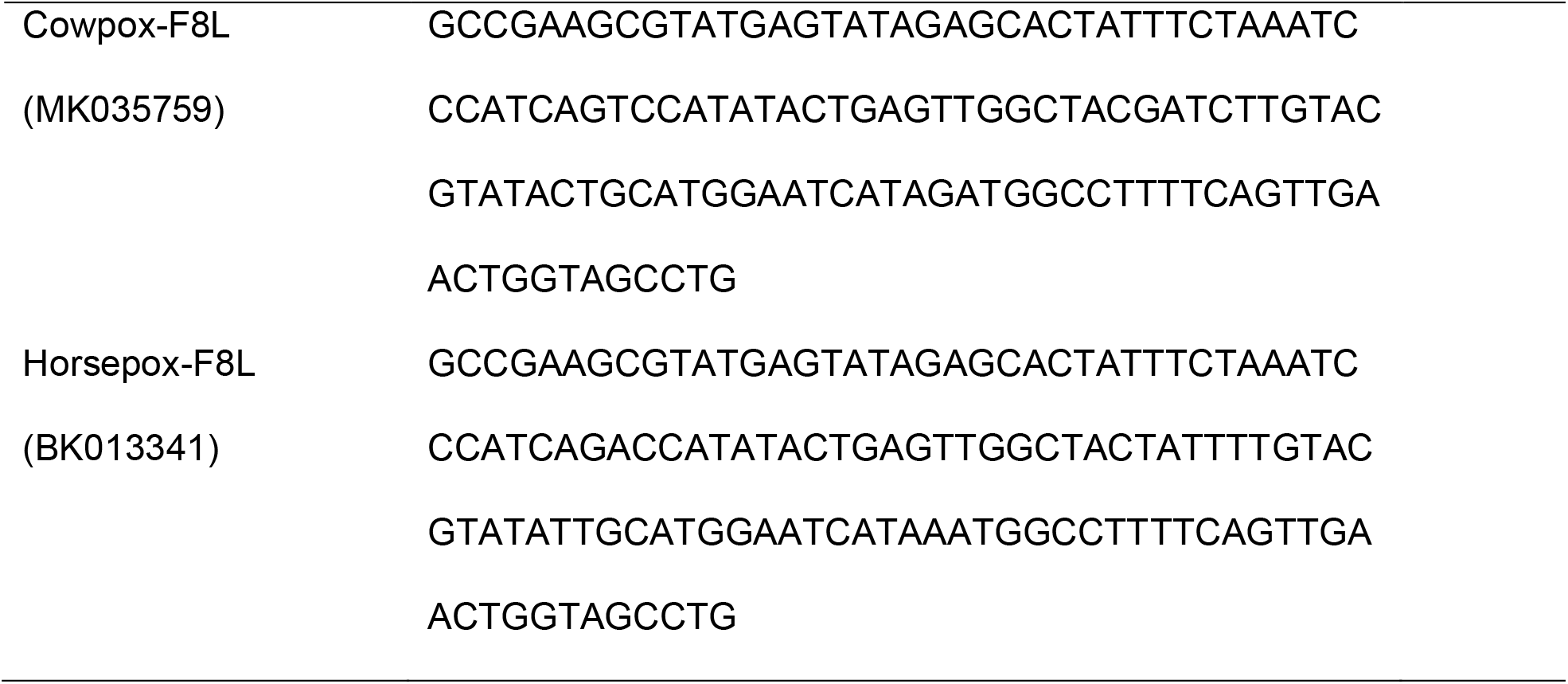
Primers, probes, and synthetic gene fragments used in this study.

